# A fly model of SCA36 reveals combinatorial neurotoxicity of hexanucleotide and dipeptide repeats

**DOI:** 10.1101/2025.05.15.25327061

**Authors:** Cheng-Tsung Hsiao, Ssu-Ju Fu, Ting-Ni Guo, Chia-Chi Lin, Yu-Jung Tsao, Yi-Chu Liao, Masayuki Hashimoto, Shu-Yi Huang, Yi-Chung Lee, Chien-Hung Yu, Chih-Chiang Chan

## Abstract

Spinocerebellar ataxia type 36 (SCA36) is a neurodegenerative disease caused by expanded (GGCCTG)n hexanucleotide repeat sequence in the *NOP56* gene. While the expanded repeats could transcribe and form toxic RNA foci within neurons, recent evidence indicates that translation of these repeats produces dipeptide repeats (DPR) that contribute to neurotoxicity. The relative impact of hexanucleotide RNA repats (HRR) and DPR on the neurodegeneration of SCA36 remains unclear. Here, we established a *Drosophila* SCA36 model to dissect the neurotoxic effects of HRR and DPR. The fly model recapitulates the cellular defects observed in SCA36 patient fibroblasts, validating its relevance for mechanistic study of SCA36. Expressing a series of the SCA36 transgenes with varying HRR lengths demonstrates an age- and length-dependent adult-onset neurodegeneration. Further engineering the transgenes to express individual DPRs reveal Proline-Glycine-DPR as the most potent neurotoxin causing progressive motor and sensory dysfunction. Interestingly, sequence modification of the transgenes to exclusively express HRR or DPR alone causes a milder phenotype, indicating both HRR and DPR contribute partially to the pathogenicity of SCA36. Therefore, this model provides a valuable platform for drug screening targeting either HRR or DPR-mediated toxicity of SCA36. Remarkably, *Spt4* knockdown or 6-azauridine treatment to suppress RNA transcription aggravates the neurodegenerative phenotypes in both the fly models and patient-derived fibroblasts, highlighting the complex interplay of pathogenic mechanisms in SCA36. These results underscore the need for carefully evaluating the potential side effects when designing therapeutic interventions for SCA36.

**Summary statement:** Spinocerebellar ataxia type 36 (SCA36) is a neurodegenerative disease caused by expanded hexanucleotide repeats. This study establishes a Drosophila model to investigate the neurotoxic mechanisms of both RNA toxicity and dipeptide repeat toxicity in SCA36. The findings highlight the complex interplay between these two factors and provide a platform for identifying potential therapeutic targets.

## Introduction

Spinocerebellar ataxia type 36 (SCA36, MIM:614153) is an autosomal dominant neurodegenerative disease caused by an expansion of a hexanucleotide repeat (GGCCTG)n (hereinafter referred to as (G3C2T)n) within the first intron of *NOP56* (NM_006392) gene. This expansion leads to various neurological symptoms, including progressive ataxia, dysarthria, sensorineural impairment, and muscle weakness, typically manifesting in adulthood of the patients (García-Murias et al., 2012; Ikeda et al., 2012; Kobayashi et al., 2011). The *NOP56* loci consists 3 to 14 repeats for unaffected individuals, while the patients affected by SCA36 carry repeats ranging from 30 to 2500 (García-Murias et al., 2012; Kobayashi et al., 2011; Lam et al., 2023; Obayashi et al., 2015). The association between the length of the repeat expansion and the clinical presentations of SCA36 patients remains to be elucidated.

Recent studies have shed light on the potential mechanisms underlying SCA36 pathology. One major hypothesis suggests that the expanded hexanucleotide RNA repeats (HRR) form toxic foci within neurons and disrupt normal cellular functions (Furuta et al., 2019; Matsuzono et al., 2017). Supporting this hypothesis, studies in cell culture showed that inhibition of RNA elongation factors, Spt4 and Spt5, could reduce RNA foci and rescue the cellular derangement (Furuta et al., 2019). However, whether inhibiting RNA elongation could lead to neuroprotection against SCA36 toxicity in a living organism remains to be determined.

Alternatively, research suggests that the expanded repeats might be translated into dipeptide repeat proteins (DPR) through upstream open reading frame (uORF)-mediated translation or non-canonical translation mechanisms. These DPR have been observed in the cells and tissues obtained from affected patients and are thought to contribute to neurotoxicity as shown in other neurological disorders of similar repeat expansion (McEachin et al., 2020; Sakae et al., 2018; Todd et al., 2020). However, a key question remains: what is the relative impact of HRR and DPR on SCA36 neurodegeneration?

To bridge the knowledge gap in SCA36 pathogenesis, we developed a model in the fruit flies *Drosophila melanogaster*. The model consists a series of modified (G3C2T)n transgenes, allowing for utilizing the versatile and powerful fly genetics to characterize the relative contribution of HRR or DPR to disease severity. The phenotypes of the SCA36 flies recapitulate the cellular defects of the fibroblasts derived from SCA36 patients. Finally, as a proof-of-principle, we utilized the fly SCA36 model to explore potential therapeutic strategies for this disease.

## Materials & Methods

### *Drosophila* strains and genetics

*Drosophila* stocks were maintained at 25°C on standard medium and genetic crosses were set up using standard fly husbandry practices. Details on fly strains and genotypes used in experiments are provided in Table 1 and on FlyBase (flybase.org), unless otherwise specified.

**Table 1.** Fly strains used in this study.

| Strains | Source | Identifier | Figure |
| --- | --- | --- | --- |
| *D. melanogaster*: UAS-(G3C2T)20 | This manuscript | N/A | Fig. 1. |
| *D. melanogaster*: UAS-(G3C2T)33 | This manuscript | N/A | Fig. 1. |
| *D. melanogaster*: UAS-(G3C2T)90 | This manuscript | N/A | Fig. 1. |
| *D. melanogaster*: UAS-(GL-DPR)33 | This manuscript | N/A | Fig. 3,4,5,7 |
| *D. melanogaster*: UAS-(AW-DPR)33 | This manuscript | N/A | Fig. 3,4,5,7 |
| *D. melanogaster*: UAS-(PG-DPR)33 | This manuscript | N/A | Fig. 3,4,5,6,7 |
| *D. melanogaster*: UAS-(AW-tandem-STOP)33 | This manuscript | N/A | Fig. 6. |
| *D. melanogaster*: UAS-(AW-codon-mod)33 | This manuscript | N/A | Fig. 6. |
| *D. melanogaster*: UAS-(PG-tandem-STOP)33 | This manuscript | N/A | Fig. 6. |
| *D. melanogaster*: UAS-(PG-codon-mod)33 | This manuscript | N/A | Fig. 6. |
| *D. melanogaster*: UAS-(AW-DPR)43-HFV | This manuscript | N/A | Fig. 3. |
| *D. melanogaster*: UAS-(PG-DPR)70-HFV | This manuscript | N/A | Fig. 3. |
| *D. melanogaster: UAS-(PG-DPR)100* | This manuscript | N/A | Fig. 3. |
| *D. melanogaster: P{UAS-mCD8:GFP.L}2* | Bloomington Drosophila Stock Center | BDSC: 5137 | Fig. 1,4. |
| *D. melanogaster: P{UAS-spt4<sup>RNAi</sup>, TRiP}* | Bloomington Drosophila Stock Center | BDSC: 31194 | Fig. 8. |
| *D. melanogaster: P{UAS-spt4<sup>RNAi</sup>, KK}* | Vienna Drosophila Resource Center | VDRC: 108459;<br>FlyBase: FBgn0028683 | Fig. 8. |
| *D. melanogaster: P{GAL4 GMR.}YH3/TM3, Sb* | Bloomington Drosophila Stock Center | BDSC: 84247 | Fig. 5. |
| *D. melanogaster: P{GAL4-elav.L}2/CyO* | Bloomington Drosophila Stock Center | BDSC: 8765 | Fig. 3,5. |
| *D. melanogaster: P{ GAL4-OK371}* | Bloomington Drosophila Stock Center | BDSC: 26160 | Fig. 1,4,6,8. |
| *D. melanogaster: P{ GAL4-71B}* | Bloomington Drosophila Stock Center | BDSC: 1747 | Fig. 7. |

### Molecular cloning & transgenesis

All cloning steps were performed according to standard protocols. Restriction enzymes and DNA ligase were purchased from New England Biolabs (NEB), while oligonucleotides and gene fragments were purchased from Integrated DNA Technologies (IDT). All of these reagents were used following the manufacturer’s sheet. The oligonucleotide sequences were listed in Table 2.

**Table 2.**
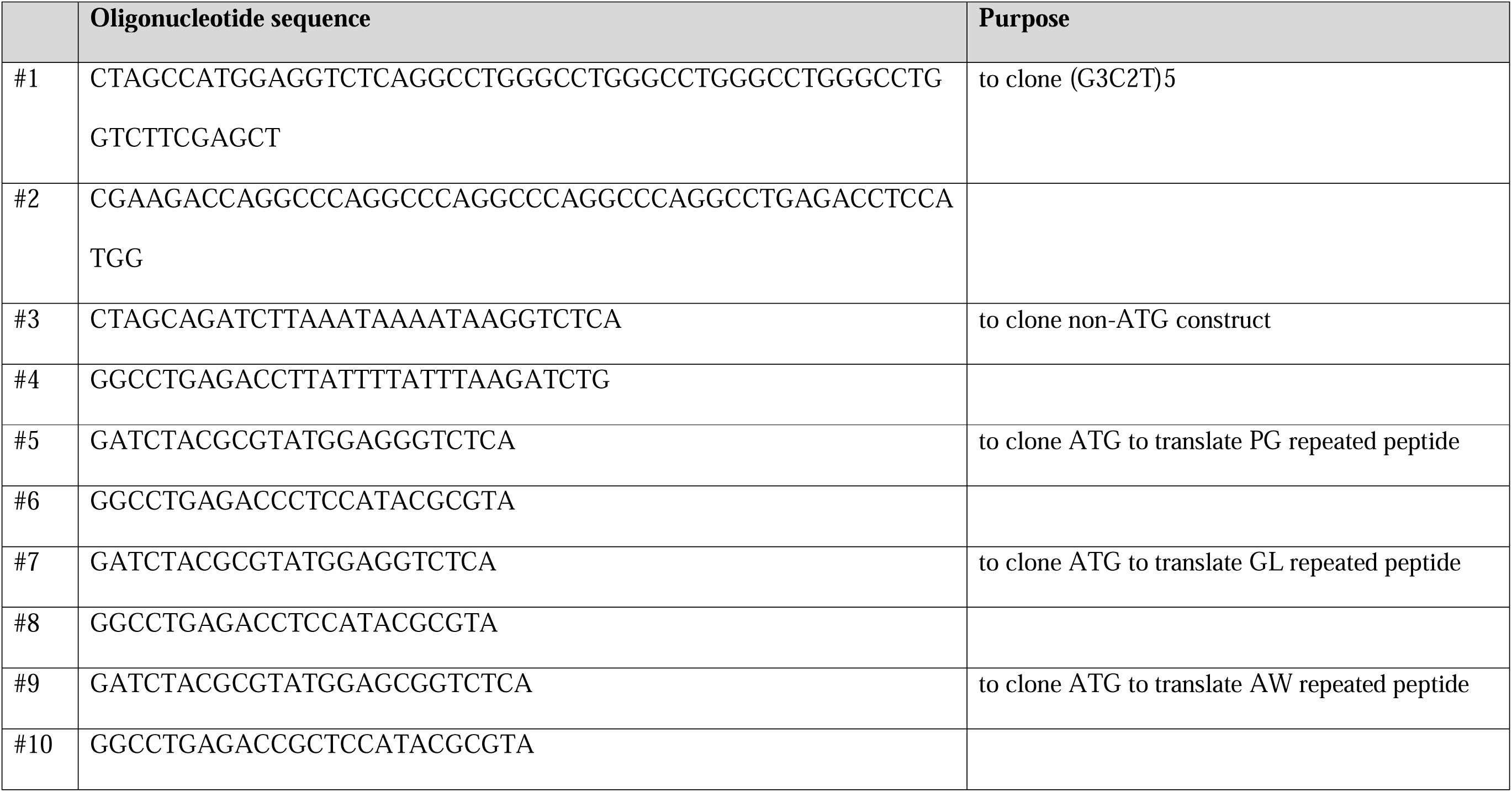
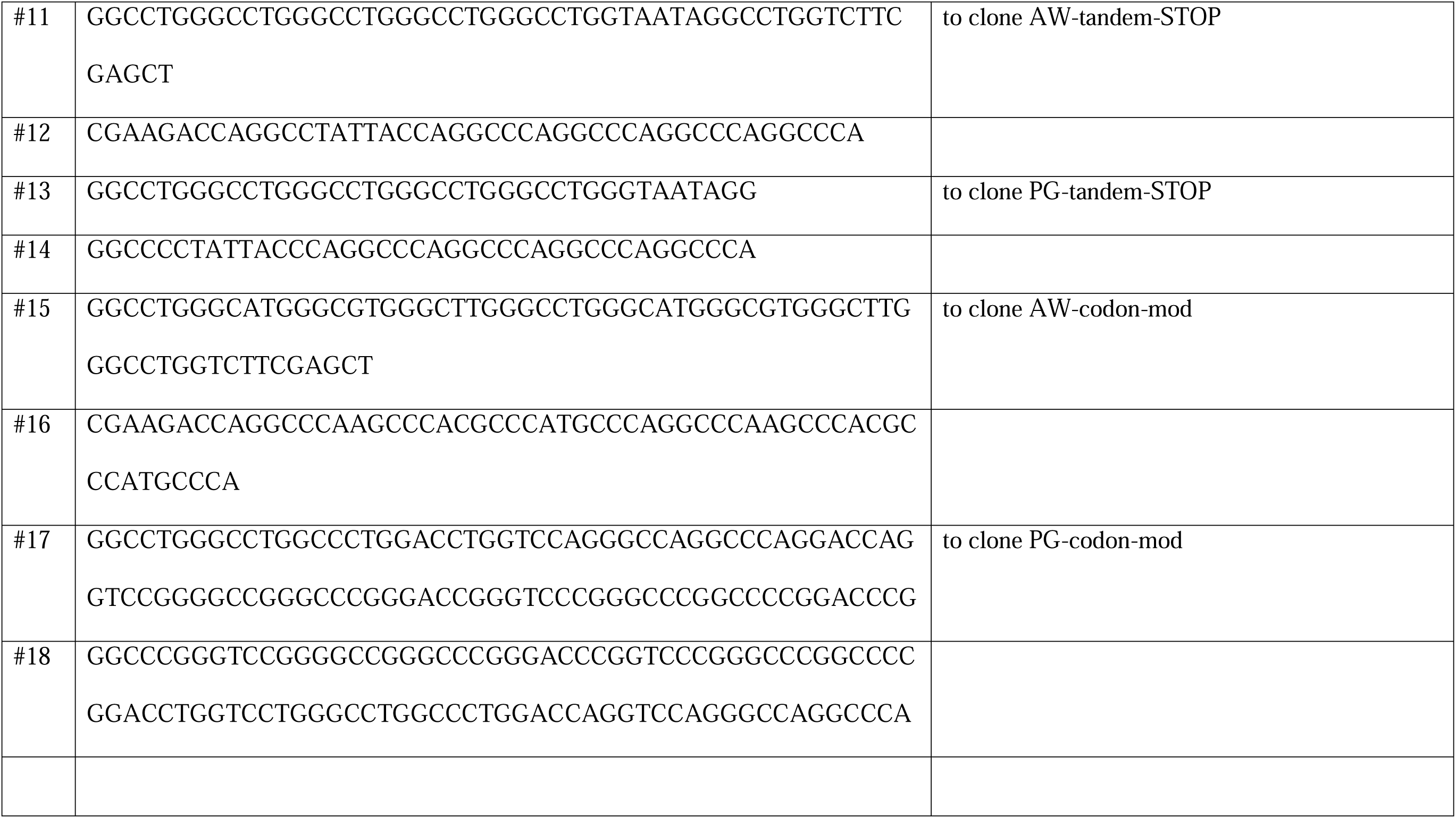
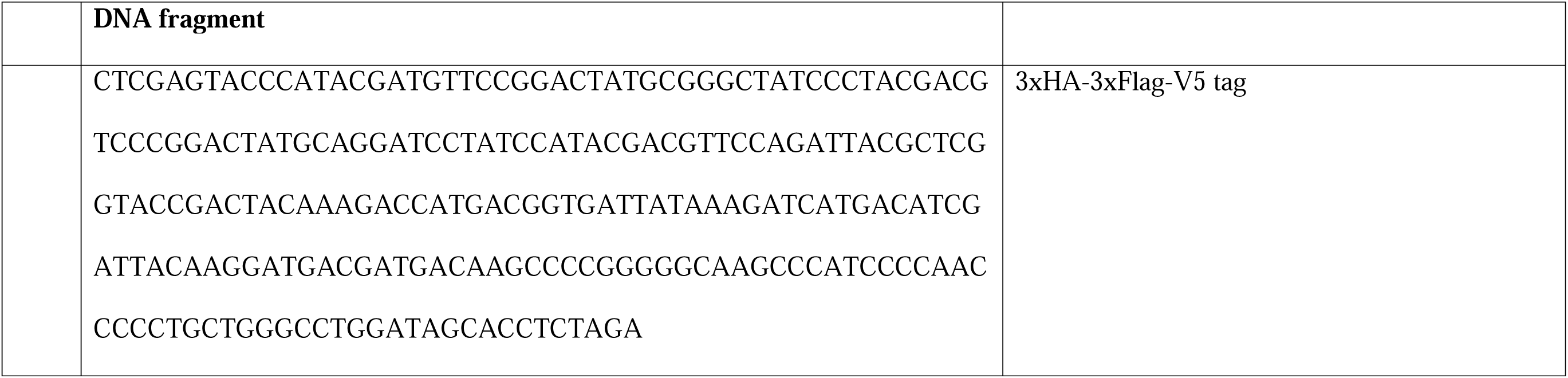
Oligonucleotides used in this study.

To construct the SCA36 (G3C2T)n repeated sequence and the associated mutants, we employed the PCR-free direct cloning method adapted from Scior *et al* (Scior et al., 2011).

The BssHII-XhoI digested fragment from pUAST-attB (Bischof et al., 2007) was first sub-cloned to pEGFP-N1 (Clontech) to obtain pEGFP-N1-pUASTattB(BX) to facilitate the following cloning process. The #1 and #2 oligonucleotides containing BsaI-5xG3C2T repeats-BbsI were annealed and then cloned into pEGFP-N1-pUASTattB(BX) via NheI and SacI to obtain pEGFP-N1-pUASTattB(BX)-(G3C2T)5. To extend the number of G3C2T repeats, the pEGFP-N1-pUASTattB(BX)-(G3C2T)5 was digested by BsaI-SacI to obtain the fragment as the insert. A parallel BbsI-SacI digestion of pEGFP-N1-pUASTattB(BX)-(G3C2T)5 generated the vector. Subsequently, the insert and vector ligation resulted in the extended G3C2T repeat numbers, e.g., pEGFP-N1-pUASTattB(BX)-(G3C2T)10. The iterative of this cloning process results in desired constructs with (G3C2T)n repeats in this study. To obtain the no-ATG constructs, the annealed #3 and #4 oligonucleotides were inserted into the designated pEGFP-N1-pUASTattB(BX)-(G3C2T)n. To obtain ATG-containing constructs, annealed #5 and #6 (for translating PG repeated peptide), #7 and #8 (for translating GL repeated peptide), and #9 and #10 (for translating AW repeated peptide) were cloned into the designated pEGFP-N1-pUASTattB(BX)-(G3C2T)n. These constructs were confirmed by Sanger sequencing.

The obtained pEGFP-N1-pUASTattB(BX)-(G3C2T)n plasmids (non-ATG) were sub-cloned to pUAST-attB via BssHII-XhoI sites for further transgenic fly generation in Fig. 1. To monitor the potential peptide products from (G3C2T)n repeats, we cloned a 3xHA-3xFlag-V5 tag (from a synthetic gene fragment) into pUATS-attB to construct pUAST-attB-HFV. Each tag is in a different reading frame to allow the identification of all possibilities of peptide synthesis of the repeated sequence, a concept similar to the previous report (Zu et al., 2011). This pUAST-attB-HFV plasmid was used as a scaffold to accommodate the (G3C2T)n repeats from pEGFP-N1-pUASTattB(BX)-(G3C2T)n through the sub-cloning via BssHII-XhoI sites. The resulting plasmids were used to generate transgenic flies, as shown in Fig. 2.

**Fig. 1.**
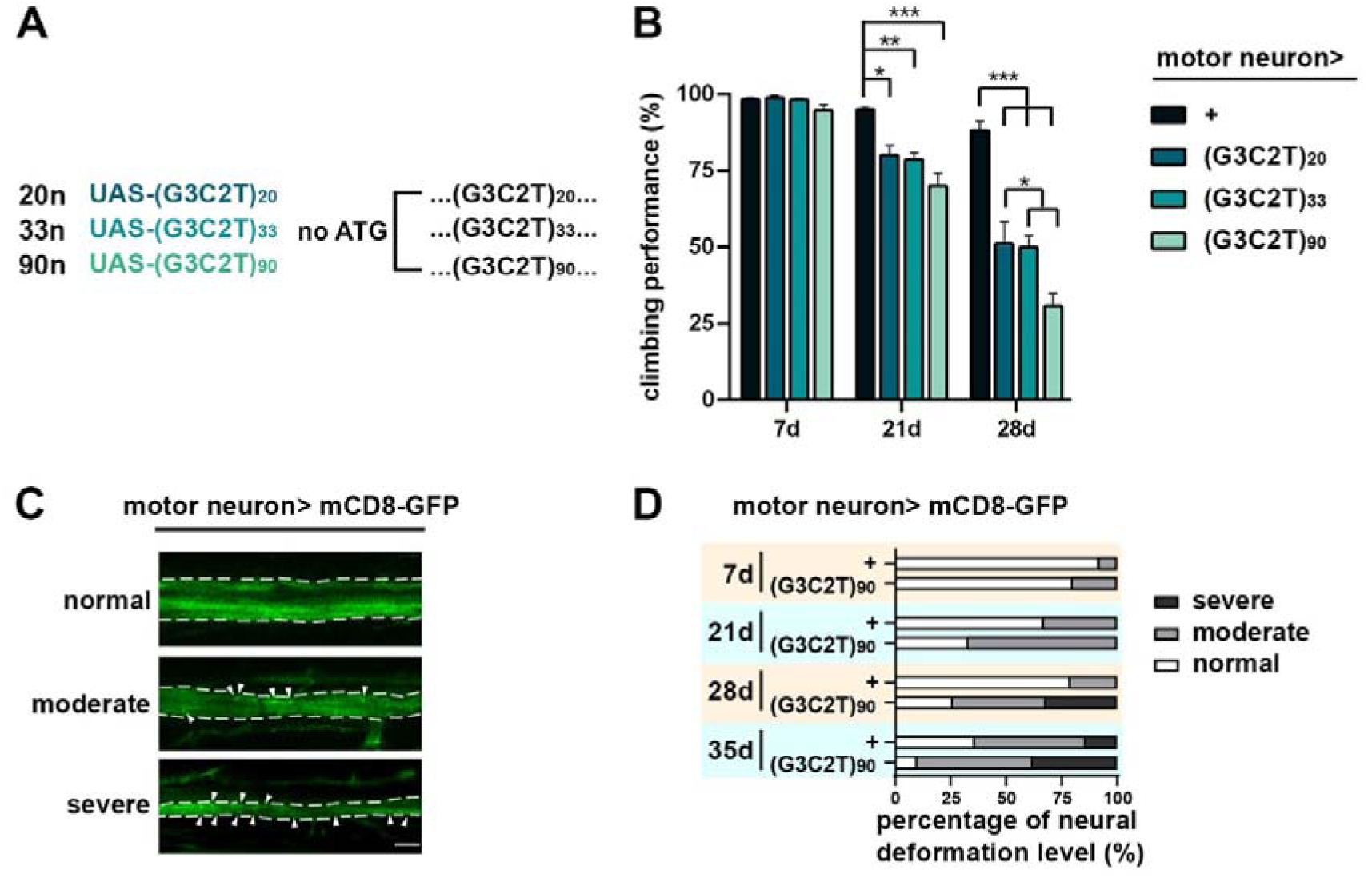
Length-associated and age-related neurodegenerative effects of the expanded (G3C2T)n transcripts. (A) Schematics of ATG-excluding transgenic constructs of UAS-(G3C2T)n of various repeat lengths, namely 20n, 33n and 90n. (B) Negative geotaxis assay of flies with transgenic expression of 20n, 33n and 90n in motor neurons. Individual UAS-(G3C2T)n hexanucleotide repeats were expressed in glutamatergic motor neuron under the control of OK371-GAL4. Flies were observed at post-eclosion day 7, 21, and 28. Number of observations: 7 independent experiments, 20-25 animals per experiment. (C) Motor axon was visualized by observing the fluorescence of the membrane-anchoring mCD8-GFP driven by OK371-GAL4. The representative confocal images of normal structure, moderated deformation, and severe deformation of motor axon were shown at the top, middle, and bottom, respectively. (Green) mCD8GFP. Arrowhead indicates the bright punctate, which are protein aggregates found in the drosophila fore femur axon. Number of observations: 7 independent experiments, 3-5 animals per experiment. Scale bar: 10 μm. (D) Quantitative analyses of axon deformation of the (G3C2T)n flies. *p<0.05, **p<0.001, ***p<0.0001, one-way ANOVA.

**Fig. 2.**
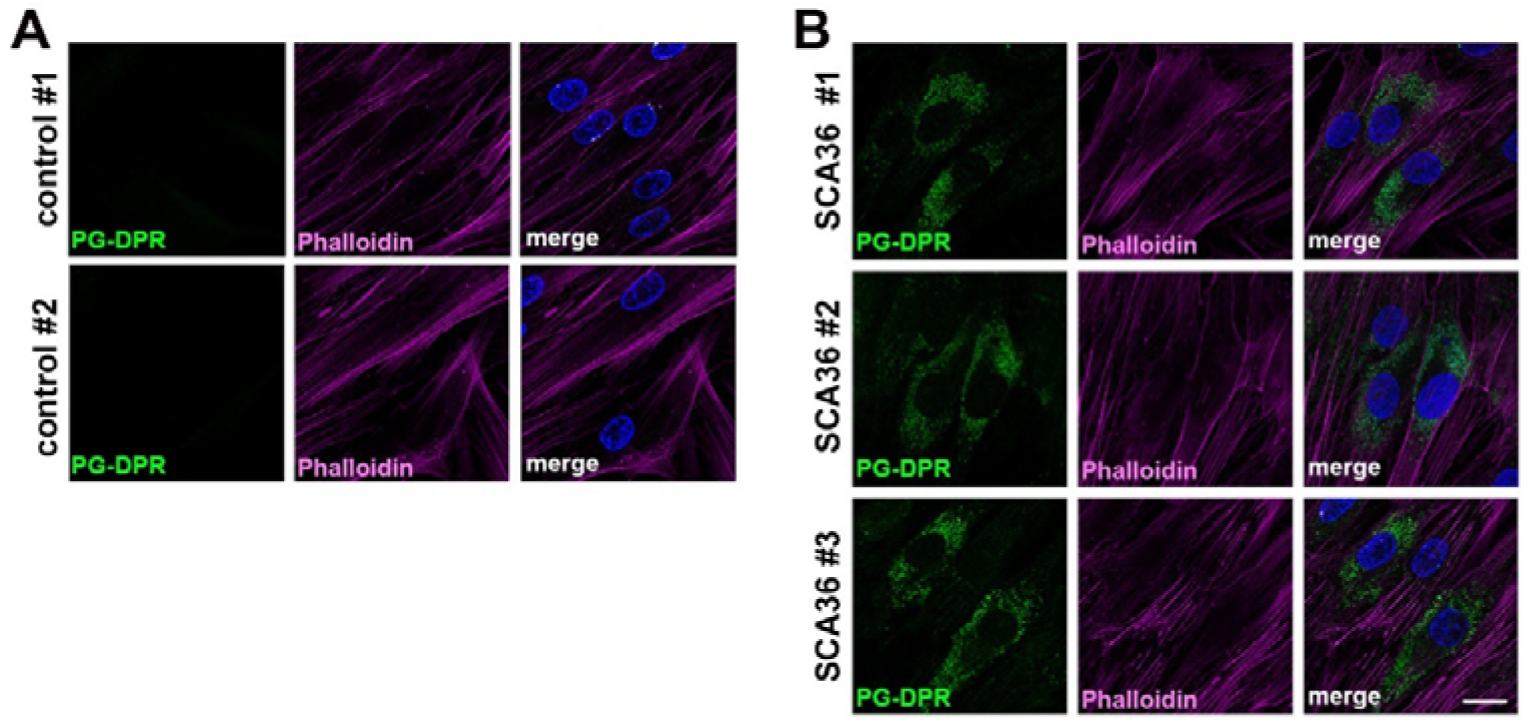
Identification of PG-DPR in the cytoplasm of patient fibroblasts. Representative immunofluorescence images showing the diffused cytoplasmic distribution without intranuclear localization of PG-DPR in fibroblasts from (A) non-patients and (B) three individual SCA36 patients. (Blue) DAPI staining to label nuclei; (Green) anti-PG immunofluorescence; (Magenta) Phalloidin staining for F-actin. Scale bar: 25 μm.

The AW-tandem-STOP and PG-tandem-STOP constructs were generated via the same strategy above, except the initial oligonucleotides are #11-#14. Similarly, the random codon modification constructs (AW-codon-mod and PG-codon-mod) were generated by the oligonucleotides #15-#18.

### Immunohistochemistry

Adult fly and larval tissues were dissected, fixed, immunostained, and mounted following our previous works (Lien et al., 2020; Tsai et al., 2020) with minor modifications on incubation and wash times. The detailed experimental conditions are described below. Information on antibodies for experiments are listed in Table 3.

**Table 3.**
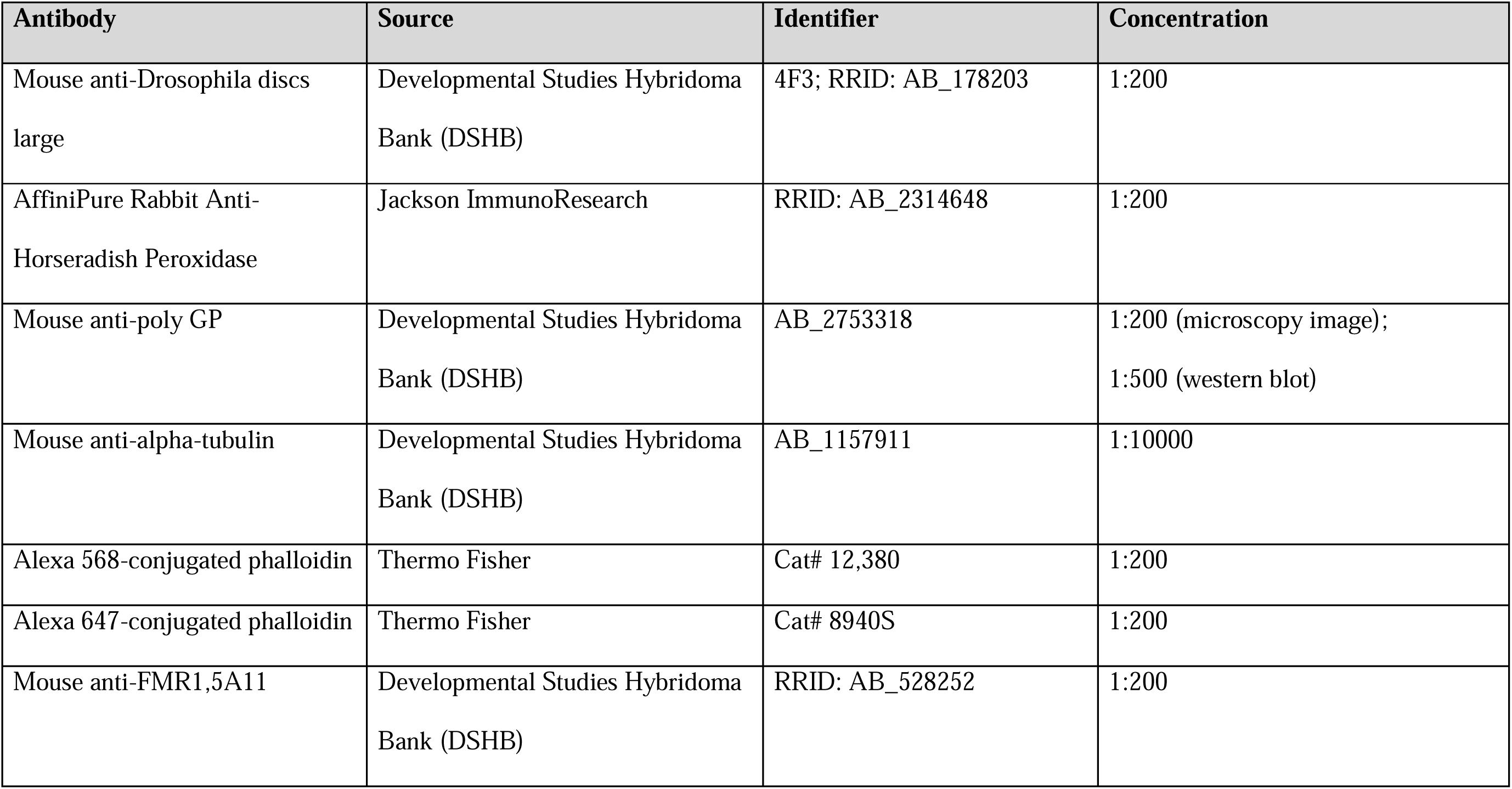
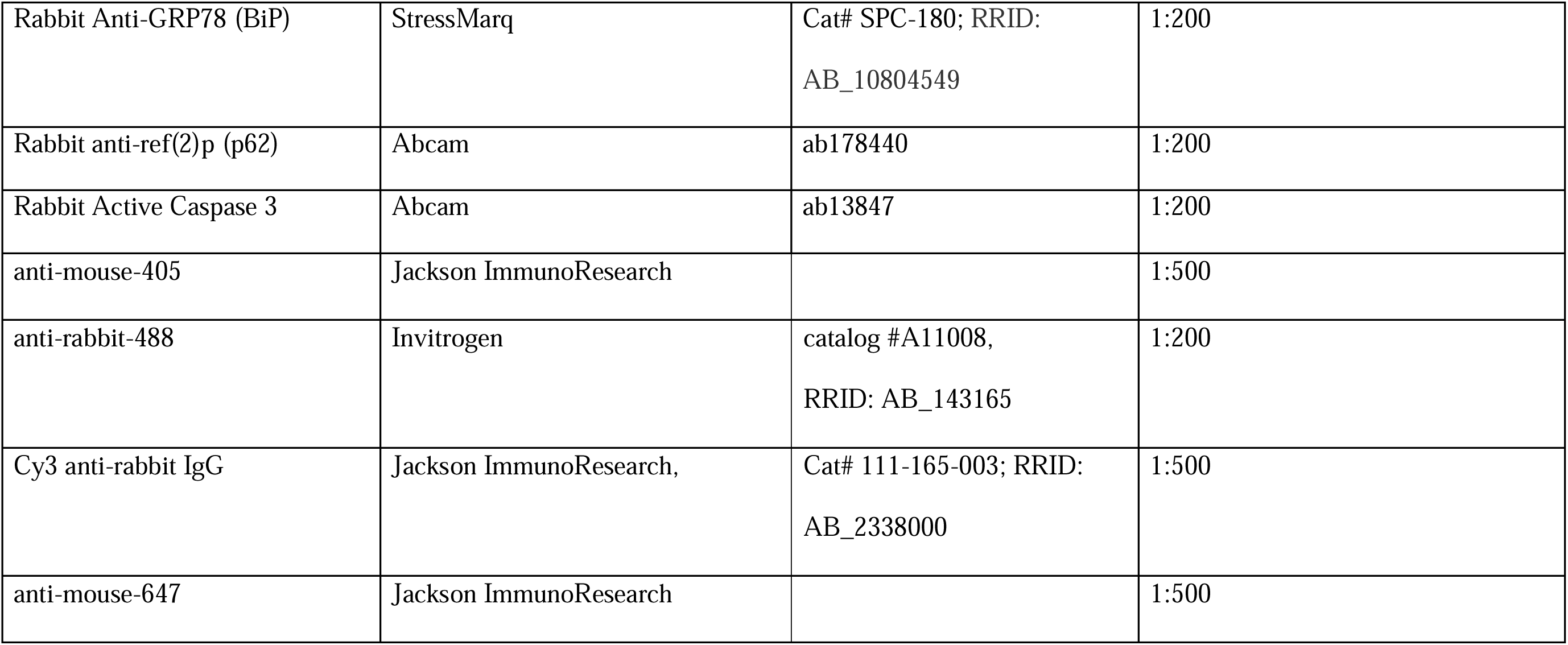
Antibodies used in this study.

### Immunofluorescence analyses for fibroblast

Fibroblast-attached coverslips were rinsed twice in 1x PBS (containing 136 mM NaCl, 2.5 mM KCl, 1.5 mM KH2PO4, and 6.5 mM Na2HPO4, pH 7.4), and then fixed with methanol on ice for 20 minutes. Fibroblasts were blocked and permeabilized with blocking buffer containing 0.1% (v/v) Triton X-100, 5% normal goat serum, 20 mM phosphate buffer, and 0.45 M NaCl, pH 7.4, for 60 minutes at 4°C. The fibroblasts were then incubated with a mouse anti-poly-PG primary antibody (1:50; Developmental Studies Hybridoma Bank) in blocking buffer overnight at 4°C. Next, fibroblasts were washed three times in blocking buffer at room temperature. Immunoreactivity was visualized with a goat anti-mouse secondary antibody conjugated to Alexa Fluor 488 (1:100; Invitrogen catalog #A11001, RRID:AB_2534069). Fibroblasts were then washed three times in 1x PBS. The cells were applied with DAPI-containing mounting medium, and fluorescence images were viewed with a Leica TCS SP8 laser-scanning confocal microscope.

### Microscopy

Images of the fly tissues including larval salivary gland, leg axon, and brain were captured with Zeiss LSM880 confocal laser scanning microscope (Zeiss microscopy, Germany). Neuromuscular junction (NMJ) and salivary gland samples were analyzed on Zeiss ApoTome.2. The human fibroblasts were imaged through Leica TCS SP8 LIGHTNING confocal microscope (Leica Camera, Germany).

### Climbing assay

The climbing assay was adapted from Hung *et al* (Hung et al., 2021). Briefly, locomotor activity was measured by counting the number of flies that climbed 7 cm in 3 seconds. Groups of 20 flies were placed in connected vials and observed for 1 minute, with 50 seconds of rest between trials. The entire process was video recorded for analysis.

### Electroretinogram (ERG)

ERGs were performed as described previously by Tzou *et al* (Tzou et al., 2021). Briefly, flies were immobilized on glass slides, and ERGs were recorded during one-second light-on and light-off cycles using electrodes filled with 2 M NaCl. Each genotype and experimental condition were tested in triplicate, with at least 10 recordings per replicate.

### Human fibroblasts

Written informed consents were provided by the participants in accordance with relevant regulatory guidelines. The study was approved by the Institutional Review Board of Taipei Veterans General Hospital. The information regarding skin biopsy acquisition were listed in Table 4. A three-millimeter punch skin biopsy was obtained from the patients by an authorized specialist. The tissues were then cut into several pieces, and the standard sample was examined by routine histopathology studies for clinical purposes. A small part of residue tissue was rinsed by 1x PBS for three times. The harvested tissue was shredded by using a dissection scissor in the culture dish. The tissue was then covered by coverslips in 20 ml DMEM culture medium supplemented with 10% fetal bovine serum (Hyclone), 1 mM sodium pyruvate, 100 units/ml penicillin, and 50 μg/ml streptomycin at 37LJ in humidified incubator with 5% CO2 and 95% air. About 20 days after the procedure, fibroblasts grown from the tissue were dissociated with 0.25% trypsin solution. The isolated fibroblasts were seeded into new culture dishes for maintenance and further experiments. The human fibroblast cells were cultured in DMEM medium (GIBCO, 11965-084) supplemented with 10% fetal bovine serum (GIBCO, A3160502) and maintained at 37°C in a 5% CO_2_ humidified incubator. The cells were passaged when reaching confluency.

**Table 4.** Fibroblast cell lines used in this study.

| Identifier | Sex of the individual | Age at the sample<br>obtained | Source |
| --- | --- | --- | --- |
| Control #1 | Female | 40's | Taipei Veterans General Hospital |
| Control #2 | Female | 50's | Taipei Veterans General Hospital |
| SCA36 #1 | Male | 50's | Taipei Veterans General Hospital |
| SCA36 #2 | Female | 50's | Taipei Veterans General Hospital |
| SCA36 #3 | Female | 40's | Taipei Veterans General Hospital |

### Western blotting for denature SDS-PAGE

Western blotting was performed as previously described (Wu et al., 2021). Protein extracted from fly heads or human fibroblast cell lines, sample lysate from tissue protein extraction reagent (Thermo, Catalog #78510) and cell lysis buffer containing the following (in mM): 150 NaCl, 5 EDTA, and 50 Tris-HCl, pH 7.6, 1% Triton X-100, 1mM PMSF, 1mM DTT and containing a protease inhibitor mixture. Protein was separated by SDS-PAGE and transferred to polyvinylidene difluoride membranes (PVDF, Millipore) as per the manufacturer’s instructions (Bio-Rad). PVDF membranes were incubated with 5% skimmed milk in TBST (10 mM Tris (pH 8.0), 150 mM NaCl, and 0.1% Tween 20) for 1 hour at room temperature, following washed with TBST for 10 minutes for 3 times, and incubated with primary antibodies at 4°C overnight. Membranes were washed three times with TBST for 10 minutes before incubation with secondary antibodies in TBST for 1 hour at RT. Blots were then washed with TBST for three times and revealed by an enhanced chemiluminescence detection system (PerkinElmer, NEL120E001EA; GeneTex, GTX14698; Thermo Scientific, 34580), and captured by the BioSpectrum™ 600 Imaging System (UVP Ltd). Quantification of western blotting results was performed using ImageJ software (RRID:SCR_003070).

### Quantification and statistical analysis

Quantitative data were analyzed using appropriate statistical tests, including two-tailed unpaired Student’s t-tests, one-way ANOVA with Tukey’s multiple comparison test, or two-way ANOVA with Bonferroni posttests. Graphs were generated using GraphPad Prism 5. Data in bar graphs are presented as mean ± SEM. Statistical significance was determined using a p-value threshold of 0.05. All statistical details are provided in the figure legends.

### AI tool usage

AI language tools, including ChatGPT and Gemini, were used to assist in the writing and editing of this manuscript. However, all data, analyses, and interpretations were conducted by the authors.

## Results

Previous studies in cultured Neuro2A cells and patient-derived iPSCs indicated that HRR of the expanded (G3C2T)n are the primary cause of neurotoxicity in SCA36 (Furuta et al., 2019; Matsuzono et al., 2017). To establish a SCA36 model in *Drosophila*, we generated a series of transgenic flies harboring the expanded (G3C2T)n, wherein transcription of the repeats is controlled by the GAL4/UAS system (Brand and Perrimon, 1993). A *Drosophila* model of FTD/ALS demonstrated that the neurotoxicity in C9orf72 correlates with the number of the expanded (G4C2)n repeats (Mizielinska et al., 2014). To examine the correlation between length of the repeated (G3C2T)n hexanucleotide and SCA36 neurotoxicity, our first set of the transgenic UAS flies was designed to carry the expanded (G3C2T)n of varying repeat lengths (hereafter referred to as 90n, 33n, and 20n, respectively) (Fig. 1A). Since this set of constructs does not contain a start codon, the potential toxicity is attributed primarily to RNA transcripts of (G3C2T)n. To examine the neurotoxic effect, we used OK371-GAL4 to specifically express the transgenes in motor neurons, followed by evaluating the neural function. Negative geotaxis serves as a behavioral assay that reflects the locomotor activity of adult flies, potentially linked to their motor function. The HRR-expressing flies exhibited normal climbing activity at day 7 post-eclosion but showed a locomotor deficit at day 21 when compared to the wild-type controls, indicating adult-onset degeneration (Fig. 1B). Notably, the deficits correlated with repeat length and age, with the 90n flies displaying more severe defects in climbing compared to those with shorter repeats at day 21 and 28 (Fig. 1B). To examine the impact on neural morphology, we marked the repeats-expressing neurons by co-expressing mCD8-GFP, a membrane-anchoring fluorescent protein, and examined the neural morphology with confocal microscopy. The morphological defect was classified from normal to severe based on bundle thickness and abnormal aggregations (Fig. 1C) (Sreedharan et al., 2015). 90n flies exhibited evident deformation of the axon bundle, and the deformation became more prominent at day 35 (Fig. 1D), indicating an age-dependent morphological degeneration. These findings indicate a length- and age-dependent correlation between the (G3C2T)n HRR and the degree of neurodegeneration.

To validate the fly model, we set to compare the findings of the fly transgenic model to both the fibroblasts we collected from SCA36 patients and existing literature. Previous literature showed that the (G4C2)n expansion of C9orf72 FTD/ALS can undergo either repeat-associated non-ATG (RAN) translation or canonical translation to produce PG-DPR. A predominant PG-DPR was found in (G3C2T)n-transfected HEK293T cells, transgenic (G3C2T)n-expressing mice, and brain tissues of postmortem SCA36 patients (McEachin et al., 2020; Todd et al., 2020). Likewise, we performed immunostaining of the SCA36 patient-derived fibroblasts to visualize PG-DPR. Immunofluorescence studies exhibited diffuse distribution of PG-DPR in the cytoplasm (Fig. 2A,B). Thus, the subcellular pattern is consistent with previous studies (Todd et al., 2020), suggesting PG-DPR is a soluble DPR. We then examined the toxicity caused by DPR in flies. For this purpose, 2 sets of transgenic flies were generated. In the first set, we inserted a tandem HA-Flag-V5 tag to the C-terminus of the (G3C2T)n expansion (Fig. 3A,B). At the 5’ end, ATG was inserted in frame with the translation of AW or PG dipeptide. This design led to in-frame translation of each DPR frame with different epitope tag, resulting in the potential production of AW-DPR-FLAG and PG-DPR-V5, respectively. The transgenes were driven by a pan-neuronal driver elav-GAL4, and subsequent Western blotting was conducted to validate the tagged DPR in fly neuron. We detected the V5-tagged PG-DPR, whereas the FLAG-tagged AW-DPR was below the detectable level (Fig. 3C). The second set was designed with an ATG start codon inserted in frame with each of the sense reading frames of (G3C2T)n, resulting in the possible translation of GL-, AW-, and PG-DPR, respectively (Fig. 3D,E). However, Western blotting of the fly lysates only revealed an accumulation of PG-DPR in the aging fly (Fig. 3F). These findings indicate the robust stability of PG-DPR, potentially suggesting its association with neurotoxic effects.

**Fig. 3.**
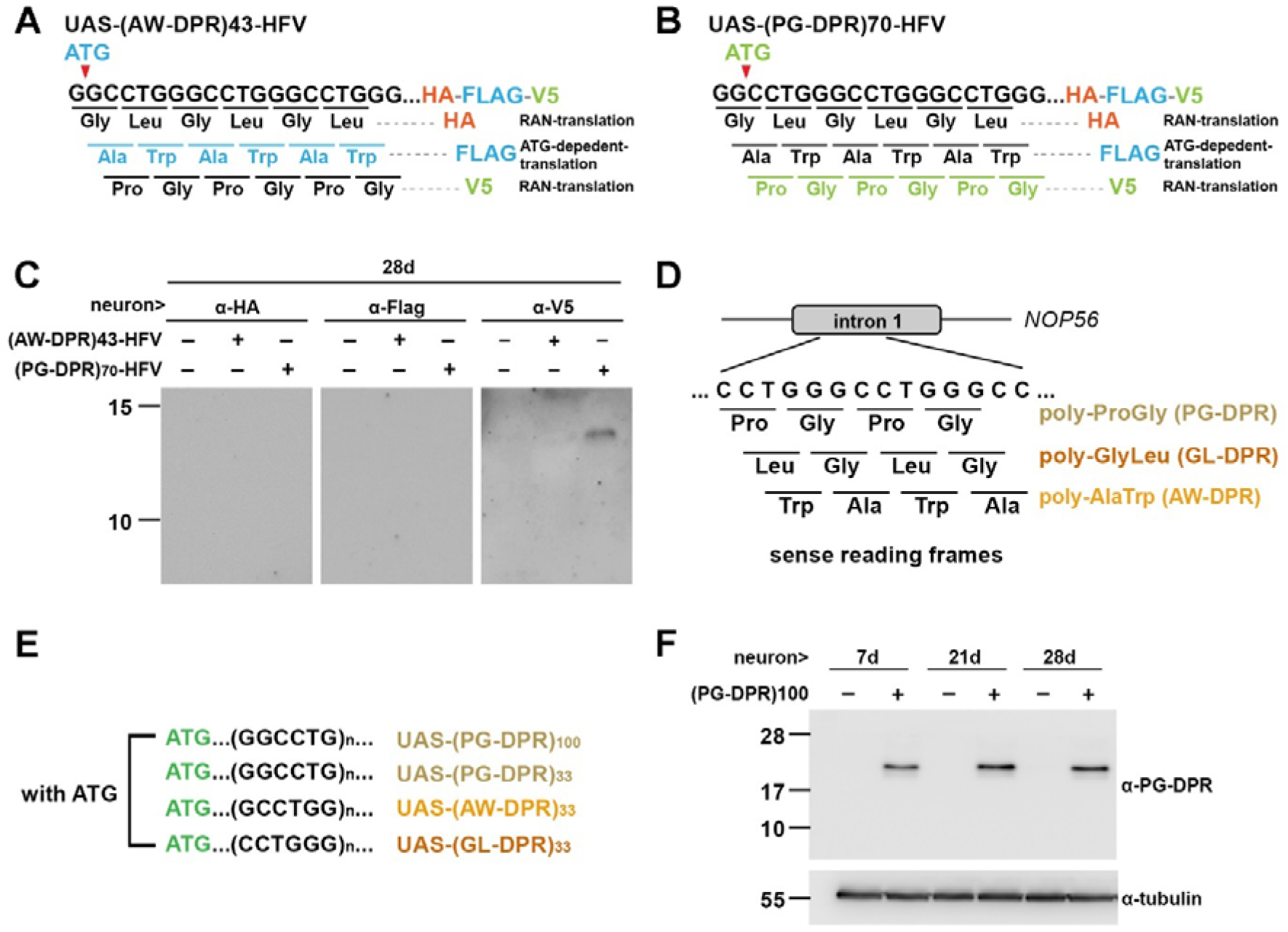
Detection of PG-DPR in adult fly brains. (A) Schematics of ATG-containing (G3C2T)43-HA-Flag-V5-tagged (HFV-tagged) constructs. These constructs were utilized to compare the effects of canonical and non-canonical translation. Of note, the ATG codon is designed in-frame with Flag-tagged AW-DPR reading frame. Whereas, HA-tagged GL-PRD and V5-tagged PG-PRD might arise from non-ATG related translation. (B) Schematics of ATG-containing (G3C2T)70-HA-Flag-V5-tagged (HFV-tagged) constructs. These constructs were utilized to compare the effects of canonical and non-canonical translation. Of note, the ATG codon is designed in-frame with V5-tagged PG-DPR reading frame. Whereas, HA-tagged GL-PRD and Flag-tagged AW-PRD might arise from non-ATG related translation. (C) Western blots of fly head extracts showing the detection of the V5-tagged PG-DPR. The HA-tagged GL-PRD and Flag-tagged AW-PRD were below the detection level. Number of observations: 5 independent experiments, 15 animals per experiment. (D) Schematics of putative DPRs translated from three reading frames of the sense expanded (G3C2T)n at *NOP56* locus. (E) Schematic presentation of putative DPRs translated from the ATG-containing (G3C2T)43 with ATG inserted at the three reading frames. (F) Representative Western blot of PG-DPR detected by anti-PG antibody in fly head extracts expressing 100 repeats of PG-DPRs driven by elav-Gal4. Flies were aged to post-eclosion day 7, 21, and 28. Number of observations: 7 independent experiments, 15 animals per experiment.

Next, we performed immunostaining of the fly tissue to investigate potential abnormalities upon PG-DPR expression. Similar to patient fibroblasts, the larval salivary glands with transgenic PG-DPR expression exhibited diffused pattern of anti-PG-DPR immunofluorescence (Fig. 4A,B,C). While PG-DPR has been shown to impair stress granule dynamics in mammalian C9orf72 model (Zhang et al., 2018), no signs of stress granule assembly was observed in poly-PG-expressing cells (Todd et al., 2020). The mixed results have prompted us to investigate the cellular impacts of each of the 3 DPR. We stained the DPR-expressing tissue with FMR1 and BiP as markers for stress granule and ER stress, respectively (Farny et al., 2009). Under confocal imaging, no obvious difference in FMR1 immunofluorescence was observed in the DPR-expressing salivary gland compared with the controls, suggesting the absence of stress granule in a fly tissue of SCA36 (Fig. 4D,E). Likewise, no difference was found when we stained with BiP as marker for ER stress, suggesting ER stress was not affected (Fig. 4 D,E). Moreover, we interrogated whether there is cellular dysregulation in autophagy and cell death by examining ref(2)p (Nezis et al., 2008) and Caspase 3 (Hung et al., 2021), respectively. No difference in immunofluorescence intensity or pattern of both markers were found in SCA36 flies compared to the controls (Fig. 4F,G), suggesting the absence of autophagy dysregulation and apoptosis activation.

**Fig. 4.**
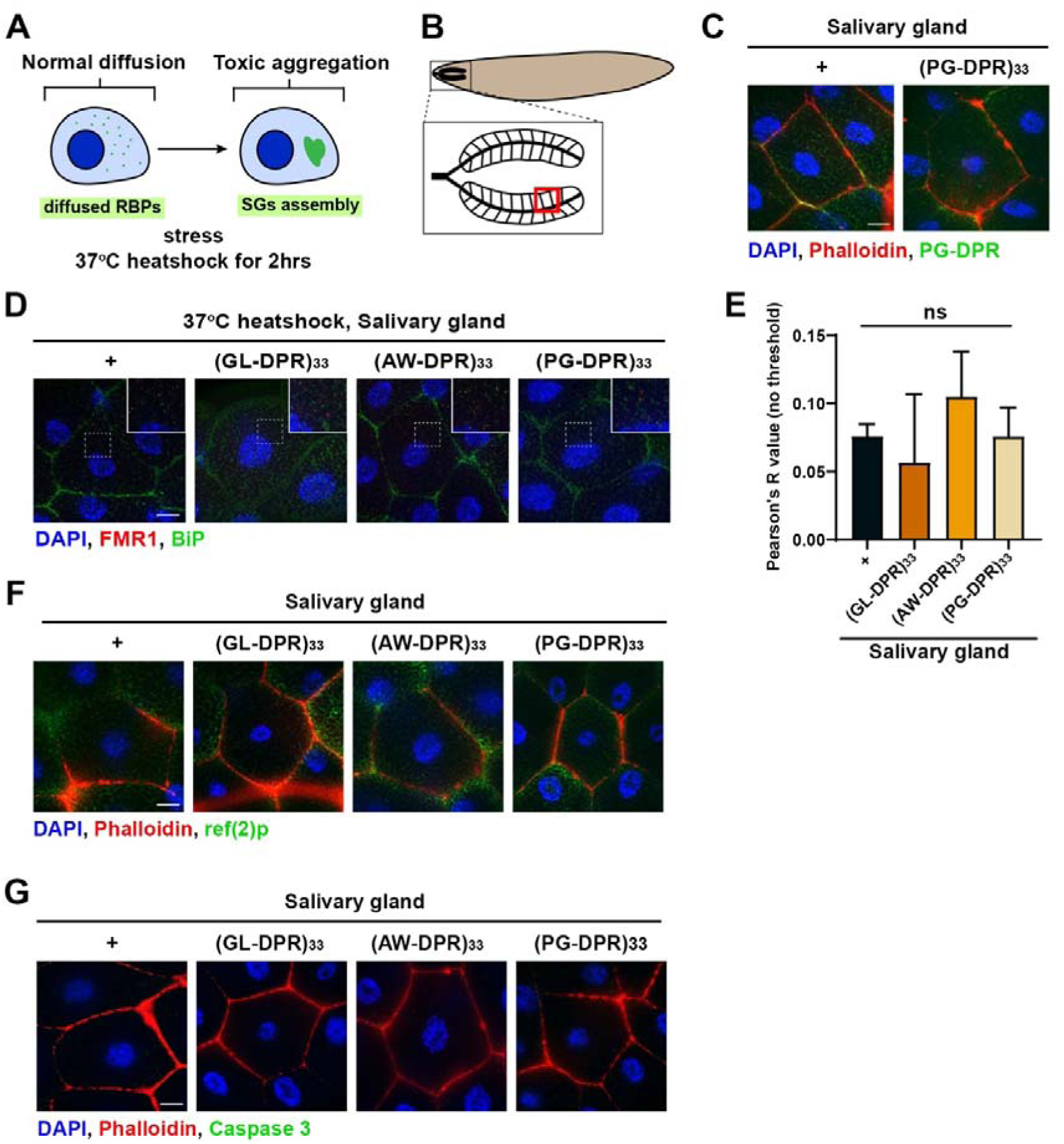
Examination of stress granule, autophagy, and apoptosis in SCA36 flies. The salivary gland 71B-Gal4 driver was utilized to express the individual DPRs in SCA36 flies. Confocal sections of salivary glands were examined to detect the presence of subtle changes related to DPRs. (A) Schematics of protein aggregation and stress granule formation. Certain types of repeat expansion disorders are susceptible to abnormal protein aggregation induced by stress stimuli, such as heat-shock induction. (B) Graphical depiction of larvae salivary glands, with a magnified region of interest indicated by the red rectangle. (C) Representative immunofluorescence microscopic images displayed the cytoplasmic distribution of PG-DPR immunoreactive to the anti-PG antibody (green). The cell membrane and nucleus were labeled with Phalloidin (red) and DAPI (blue), respectively. Number of observations: 10-12. Scale bar: 20 μm. (D) Immunofluorescence images delineated the intracellular localization of anti-fragile X messenger ribonucleoprotein 1 (anti-FMR1) antibody (red) to identify stress granules and anti-BiP antibody (green) to detect endoplasmic reticulum (ER) stress in the presence of three individual GL-, AW-, and PG-DPR, as compared to the control. Number of observations: 10-12. Scale bar: 20 μm. (E) Quantification of the immunofluorescence images using Pearson’s coefficient analysis, which demonstrated no significant differences between each group. Number of observations: 6-8. (F) Microscopic images showed the salivary glands stained with anti-ref(2)p antibody (green), indicating no significant activation of autophagy in flies expressing the three individual DPRs, as compared to the controls. Number of observations: 10-12. Scale bar: 20 μm. (G) Microscopic images showed the salivary glands stained with anti-Caspase 3 antibody (green), indicating the absence of enhanced apoptotic cell death in the flies expressing the three individual DPRs, as compared to the control flies. Scale bar: 20 μm. Number of observations: ≥15.

We then investigated the neurotoxic effects of the DPR by confocal imaging. Transgenic DPR expression in motor neurons led to age-dependent degeneration, as revealed by noticeable neural deformation and aggregation within the axon bundle (Fig. 5A,B). Among the 3 DPRs, PG-DPR exerted the most pronounced toxicity, evident from severe axon deformities compared to the other 2 DPRs upon aging (Fig. 5A,B). Similarly, flies expressing PG-DPR in motor neuron showed a more rapid deterioration in locomotor activity when comparing to the other 2 DPRs, indicating functional impairments in SCA36 fly models might be predominantly associated with PG-DPR (Fig. 5C).

**Fig. 5.**
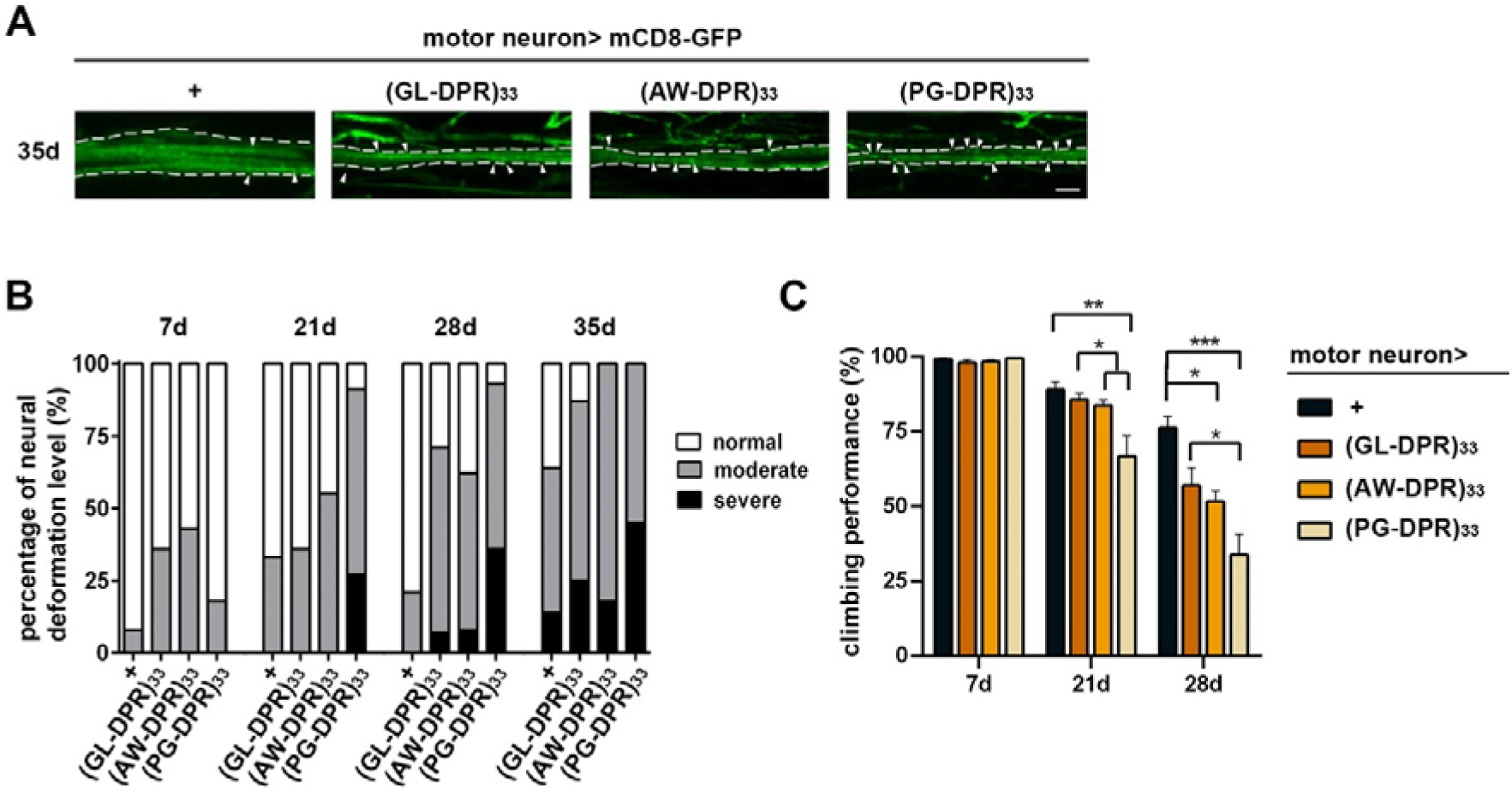
Expression of PG-DPR causes severe neurodegenerative effects. Alternate reading frames of the ATG-containing hexanucleotide repeats corresponding to GL-, AW-, and PG-DPR of equal expansion repeats were respectively expressed to compare the toxic effects of each DPRs. (A) Representative confocal images of axon bundles of the post-eclosion day 35 flies expressing (DPRs)33, showing age-dependent axon deformation. Scale bar: 10 μm. (B) Quantitative analyses of axon deformation of flies expressing individual DPRs at day 7, 21, 28, 35 post-eclosion. Number of observations: 5 independent experiments, 3-5 animals per experiment. (C) Negative geotaxis analyses of DPRs-expressing flies demonstrated age-associated declines of climbing performance at day 21 and 28 post-eclosion, with most severe phenotype observed in flies expressing PG-DPR at day 28. Number of observations: 10-12 independent experiments, 15-20 animals per experiment. *p<0.05, **p<0.001, ***p<0.0001, one-way ANOVA.

SCA36 is considered a late-onset neurodegenerative disorder, typically affecting the central nervous system in adulthood (García-Murias et al., 2012; Ikeda et al., 2012; Kobayashi et al., 2011). Therefore, we used the adult fly brain as a test tube to examine the DPR impact. The vacuolization of the CNS resulted from neuronal loss were visualized by counterstaining of DAPI. PG-DPR-expressing flies exhibited significant vacuolation compared with wildtype controls at day 45 post-eclosing, indicating the degenerating brain (Fig. 6A,B). Sensorineural impairment has been observed in SCA36 patients (García-Murias et al., 2012; Ikeda et al., 2012). Therefore, we asked whether expressing DPR may also lead to functional deterioration in fly sensory system. To achieve this goal, individual DPR were expressed specifically in the fly photoreceptor sensory neurons using the GMR-Gal4 driver. The photoreceptor function was assessed with electroretinogram (ERG). The amplitude of receptor potential serves as a readout for photoreceptor health, as the decrease in amplitude indicates neurodegeneration. The PG-DPR and AW-DPR groups exhibited a greater reduction in receptor potential compared to the GL-DPR group at day 21, suggesting the former two groups having a more pronounced toxic effect on the aging photoreceptors (Fig. 6C,D). Taken together, our fly model demonstrated the significant involvement of DPR-related toxicity in the pathogenesis of SCA36, with PG-DPR exhibiting the most notable neurotoxicity in the CNS as well as the sensory neurons.

**Fig. 6.**
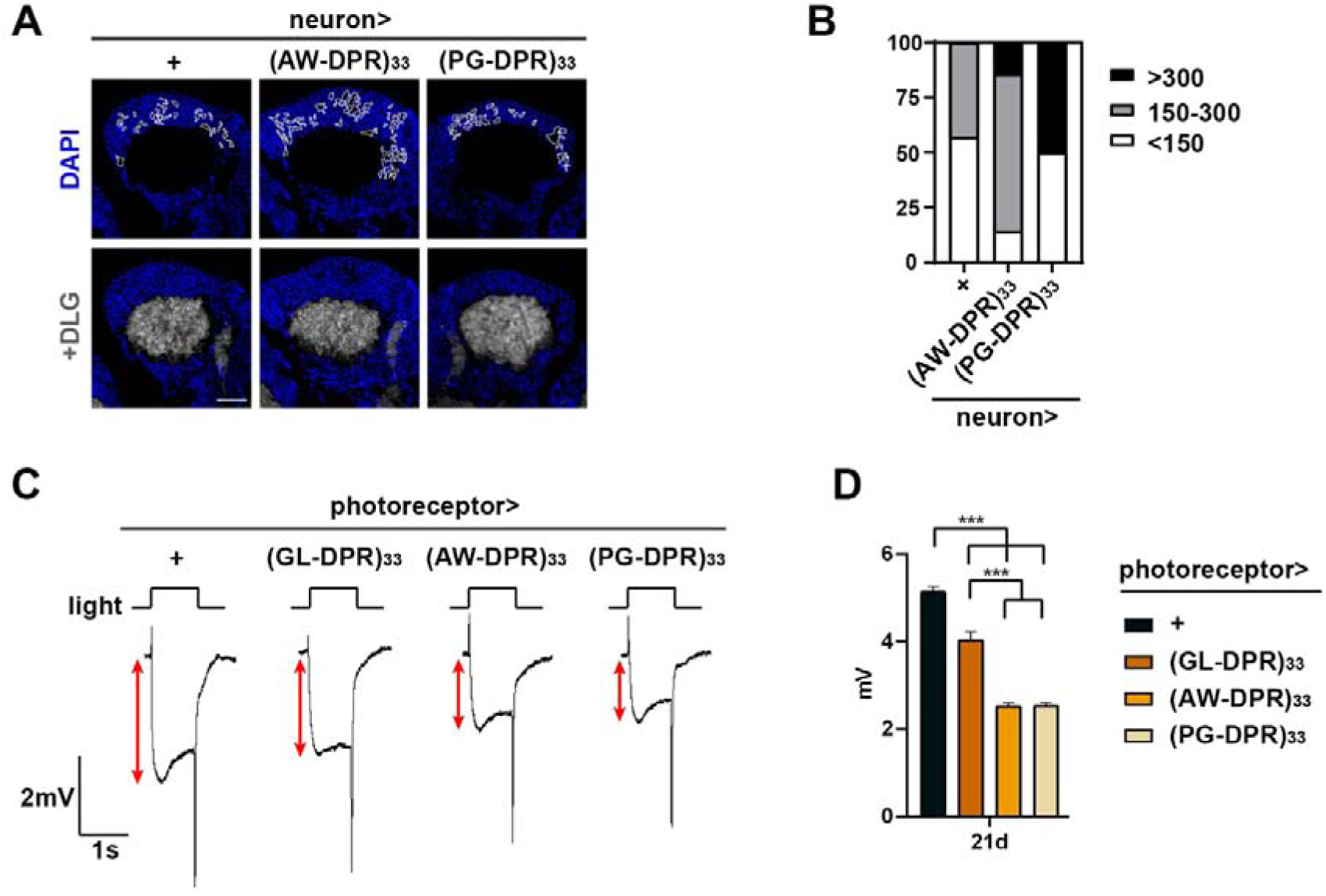
DPR-expressing flies display neuronal loss and attenuation of neuronal activities. Transgenic expression of individual DPRs in the flies photoreceptor and brain demonstrated the deteriorating effects from the DPRs. (A) Brain vacuoles indicated by absence of DAPI staining in the fly brain at post-eclosion day 45. PG-DPR-expressing brains exhibited the biggest sum of vacuole areas, indicating most prominent neuronal loss. Scale bar: 20 μm. (B) Quantification of total vacuoles area in (A). The total area >300 μm^2^, 150-300 μm^2^, and <150 μm^2^ were referred to the severe, moderate, and mild, respectively. (C) Electroretinogram (ERG) responses during photo stimulation to an 1-sec pulse of white light. Representative ERG traces of the control and three individual GL-, AW-, and PG-DPRs under GMR-Gal4 driver at post-eclosion day 21. The red double arrow indicated the amplitudes of receptor potentials. (D) Quantification of the receptor potentials in (C). Number of observations: 3 independent experiments, 8-11 animals per experiment.

While our findings and others suggest that neurotoxicity in SCA36 patients can be attributed to DPR, the impact of HRR may also exert additional impact on SCA36 pathogenesis as demonstrated in Fig. 1 and observed in previous studies (Furuta et al., 2019; Matsuzono et al., 2017). To pinpoint the primary cause of neurotoxicity, we engineered a set of constructs to distinguish between toxicity arising from HRR and that stemming from DPR (Fig. 7A,B). The PG-tandem-STOP construct featured a stop codon inserted in every 5-6 dipeptide repeats, effectively disrupting the formation of extended DPR (Fig. 6A). Conversely, the PG-codon-modified construct altered the third amino acid of each codon, thereby disrupting the repetitive nature of HRR while preserving the repetition of the DPR (Fig. 7B). Expression of either the tandem-STOP or codon-modified transgenes in neurons caused locomotor decline, suggesting that both HRR and DPR contribute to the functional impairment in SCA36 (Fig. 7C). Notably, while both transgenes caused significant locomotor decline, the deterioration was not as severe as the PG-DPR transgene alone which produces both HRR and DPR concomitantly (Fig. 7C). Taken together, our findings demonstrate a combinatorial toxicity involving HRR and DPR underlying the pathogenesis of SCA36.

**Fig. 7.**
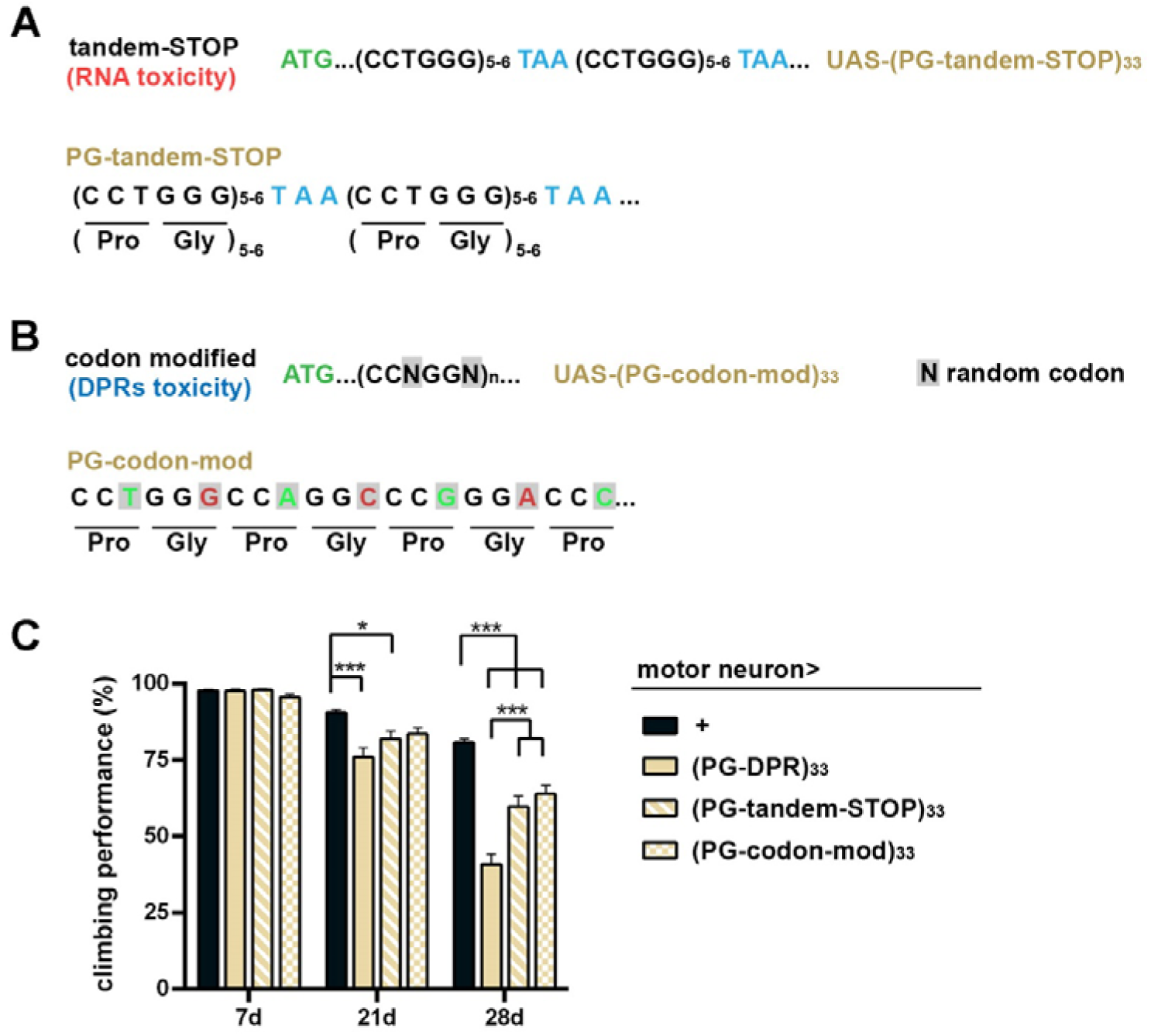
Combinatorial neurodegenerative effects from DPR and (G3C2T)n transcripts in SCA36 flies. (A) Schematics of PG-tandem-STOP construct. STOP codons were inserted for translational termination. (B) Schematics of PG-codon-modified construct. The third nucleotide of each codon was modified to disrupt the repetition at DNA/RNA level but reserve the amino acid repetition of PG-DPR. (C) Climbing assay to assess locomotor deficit related to the toxic effects of PG-DPR, PG-tandem-STOP, and PG-codon-modified constructs. Number of observations: 7 independent experiments, 15-20 animals per experiment. *p<0.05, **p<0.001, ***p<0.0001, one-way ANOVA.

Finally, we interrogated whether the transcription elongation machinery, a potential therapeutic strategy for repeat expansion disorders, would ameliorate SCA36 neurotoxicity. Although knockdown of *Spt4* leading to a reduction in RNA foci has shown beneficial effects in cellular models (Furuta et al., 2019), its therapeutic effect has never been tested in a living organism. We assessed the functional impact of *Spt4* knockdown in our fly SCA36 model and patient-derived fibroblasts. Surprisingly, knockdown of *Spt4* in the motor neuron using 2 independent RNAi strains was not able to ameliorate the locomotor defects of (G3C2T)n-induced neurotoxicity. Instead, it paradoxically exacerbated the phenotypic severity (Fig. 8A,B). Similarly, treatment with 6-azauridine (6-AZA), a small molecule that potentially targets the Spt4/5 interaction, also aggravated these defects (Fig. 8C). Furthermore, 6-AZA treatment to the fibroblasts induced stress granule accumulation in a dose-dependent manner (Fig. 8D). These findings suggest that suppression of RNA elongation may not alleviate the locomotor defect of SCA36 flies, indicating that (G3C2T)n-driven neurodegeneration may proceed via pathways beyond the Spt4-dependent mechanisms.

**Fig. 8.**
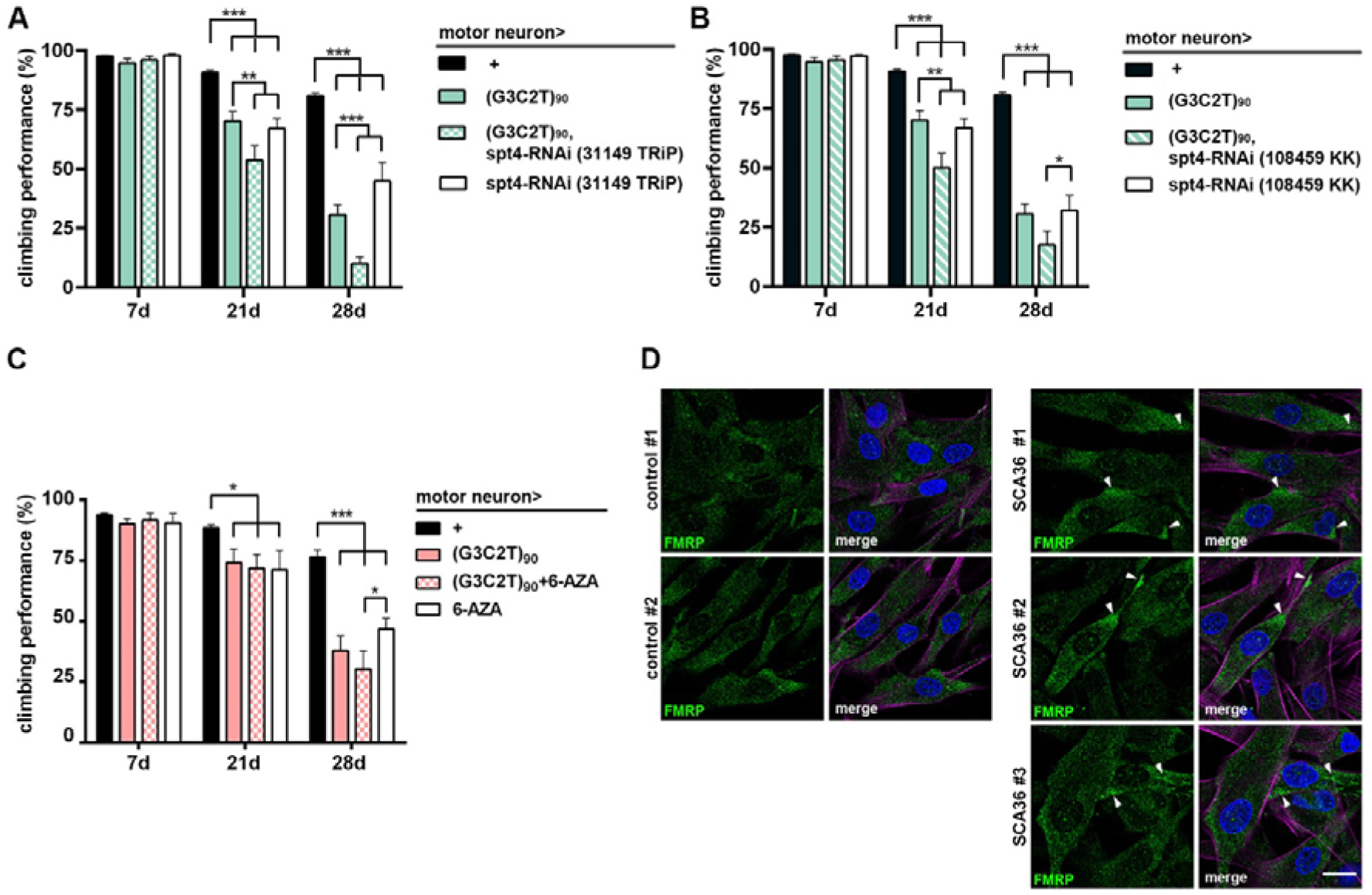
Knockdown of *Spt4* resulted in locomotor impairment of SCA36 flies. (A and B) Two independent *Spt4*-RNAi, driven by OK371-Gal4, were utilized to knockdown *Spt4* in glutamatergic neurons of flies. The climbing abilities of adult flies were examined at post-eclosion day 7, 21, and 28. The flies expressing (G3C2T)90 with *Spt4*-RNAi exhibited the most severe locomotor deficit compared to the (G3C2T)90, *Spt4*-RNAi, and control counterparts at post-eclosion day 21 and 28. (C) Treatment of 6-azauridine (6-AZA) from the larva stage to the given day of experiments. Negative geotaxis assays were evaluated at day 7, 21, and 28. Number of experiments: 7-9 independent experiments, 15-20 animals per experiment. *p<0.05, **p<0.001, ***p<0.0001, one-way ANOVA. (D) Administration of 6-AZA in fibroblasts from 3 independent SCA36 patients. FMRP immunoreactive signals were observed by confocal microscopy to examine stress granule formation. Arrowheads indicated aggregated FMRP signals. Number of experiments: 3 independent experiments.

## Discussion

In this study, we characterized the neurotoxic effects of a series of modified (G3C2T)n repeat expansion in *Drosophila* to reveal the molecular pathogenesis of SCA36. Our experiments demonstrated a positive correlation between the repeat length and severity of neurodegeneration (Fig. 1). Also, our data demonstrated that both HRR and DPR contribute to neurotoxicity and serve as critical factors in the molecular pathogenesis of SCA36. Among the different DPR products, PG-DPR exerted the most potent toxic effects (Fig. 5,6). The tandem-STOP and codon-modified transgenes further confirmed the combinatorial effects of HRR and DPR on the neurotoxicity that led to SCA36 neurodegeneration (Fig. 7). Taken together, the fly model of SCA36 recapitulates several key aspects of the human disorder, including locomotion disability, sensory neuron injury, and late adulthood onset disease. Our study provides a valuable tool for understanding the molecular mechanisms and a drug screening platform for potential upcoming therapies.

Unraveling the disease-underlying mechanisms of non-coding repeat expansions remains a significant challenge. The HRR-associated toxicity is considered as a crucial factor for repeat expansions disorders, since previous research identified the neurotoxic RNA foci in models of C9-ALS-(G4C2)n and SCA36-(G3C2T)n. Interestingly, while large RNA aggregation foci were observed in C9-ALS, SCA36 is characterized by smaller RNA aggregates (McEachin et al., 2020; Todd et al., 2020). Our series of transgenic flies carrying different repeat lengths demonstrated a positive correlation between the length of (G3C2T)n HRR and severity of neurotoxicity (Fig. 1). Upon introducing these various repeat lengths in the motor neuron, significant neurodegeneration was evident (Fig. 1C,D). This observation validated the HRR toxicity in motor neurons.

In terms of DPR toxicity, the (G3C2T)n repeat within the sense reading frame of *NOP56* potentially could generate three kinds of DPR: PG, AW, and GL, raising the questions regarding the individual contributions of each DPR to the disease pathogenesis. However, studies in C9-ALS demonstrated that selective DPR reduction did not yield clinical benefit (van den Berg et al., 2024), highlighting the disease complexity and supporting the necessity of examining the pathogenic characteristics of each DPR. The sole presence of PG-DPR in SCA36 was attributed to the uORF-mediated translation (McEachin et al., 2020). Furthermore, as introducing the PG-DPR in the fruit flies, the PG-DPR appeared to be diffusely distributed in the cytoplasm (Fig. 4C). Similarly, a previous study had also suggested GP-DPR do not aggregate in SCA36 patient tissue (McEachin et al., 2020). In contrast to DPR in C9-ALS, which require combination of specific DPR for aggregation. Elucidating the toxic mechanisms from individual DPR awaits further studies for SCA36.

To simultaneously evaluate the relative toxicity raised from individual DPR, we compared the effects of specific DPR in flies. The flies expressing the GL-, AW-, PG-DRPs displayed obvious neurodegenerative phenotypes in the brain, motor neuron, and sensory neurons (Fig. 5,6). This age-associated neurodegeneration is most prominent when expressing the PG-DPR. Despite 33n DPR being efficient to induce neurotoxic effects, they appeared to contribute less in disrupting the stress granule formation, ER-mediated apoptosis, and autophagy flux (Fig. 4). It remains an open question whether these effects would be similarly observed in the patient samples and in the animals expressing the longer DPR, or if the presence of additional contributing factors is required. Nonetheless, the partial neurotoxicity of tandem-STOP and codon-modified transgenes suggests the combinatorial effect of HRR and DPR exerts the most significant impact on the pathophysiological changes associated with SCA36 (Fig. 7). These findings provide valuable insights into the complexity of the pathogenesis of SCA36, demonstrating that our multimodal approach could be utilized to investigate the therapeutic effects of potential drug targets.

Spt4 and Spt5 form a complex that facilitates RNA polymerase II in transcription elongation (Uzun et al., 2021), thus regulates HRR transcription and DPR translation. Because the Spt4/5 complex may stabilize the transcription of extended tandem repeats, Spt4 inhibition has been postulated as a potential therapeutic target for repeat expansion disorders. In fact, inhibiting Spt4 has been shown to reduce poly-GP HRR and DPR expression in animal models and iPSC-derived cortical neurons of C9-ALS-(G4C2)n (Kramer et al., 2016). Similarly, in models of Huntington’s disease, Spt4 depletion selectively reduced trinucleotide repeats mRNA and polyQ mutant huntingtin protein levels (Liu et al., 2012). To assess whether disrupting Spt4 could be a viable therapeutic approach, we examined the effects of *Spt4* knockdown on neuronal function in the fly SCA36 model. Surprisingly, Spt4 inhibition not only failed to restore the locomotor deficits but exacerbated the neurotoxic effects (Fig. 8A,B). Instead of rescuing locomotor function, administration of 6-AZA, an inhibitor of the Spt4/5 complex, worsened the phenotype upon aging (Fig. 8C). Moreover, treatment with 6-AZA in patient-derived fibroblasts resulted in marked accumulation of stress granules (Fig. 8D). These findings suggest that HRR and DPR toxicity may involve mechanisms beyond the Spt4-dependent pathway, leading to unanticipated toxicity. Our results emphasize the need for further research to clarify these mechanisms and guide the development of targeted therapeutic strategies.

In conclusion, we have developed a *Drosophila* SCA36 model displaying cellular and phenotypical deficit resemble the observations in patients. Our experiments have demonstrated the synergistic effects of HRR and DPR to the complexity of SCA36 pathogenesis. Furthermore, the SCA36 flies and the patient-derived fibroblasts can be utilized to validate the potential therapeutic strategies for SCA36.

## Data Availability

All data produced in the present study are available upon reasonable request to the authors

## Acknowledgements

We are grateful to the patients’ participating in this study. We thank the Bloomington Drosophila Stock Center and the Vienna Drosophila Resource Center for providing the fly strains used in this study. We are grateful to WellGenetics Inc. for the technical support in generating fly strains. We thank the Developmental Studies Hybridoma Bank for the antibodies. We thank the staff of the Biomedical Resource Core and Imaging Cores at the First Core Labs, National Taiwan University College of Medicine, for excellent technical assistance in confocal imaging. This work was supported by grants from the National Science and Technology Council of Taiwan (NSTC 112-2314-B-075-039- and 113-2314-B-075-073-MY3 to C.-T.H.; 111-2320-B-002-049-MY3, 112-2314-B-002-016, and 113-2320-B002-022-MY3 to C.-C.C.; 113-2636-B-006-006 to C.-H.Y.), National Health Research Institutes (EX112-11228NI and EX113-11228NI to C.-C.C.), and National Taiwan University (112L895403 and 113L893103 to C.-C.C.).

## Data and resource availability

The datasets generated and analyzed in this study are available from the corresponding author upon reasonable request.

## Author contributions

Conceptualization: C.-T.H., S.-J.F., T.-N.G., C.-H.Y., C.-C.C.; Methodology: S.-J.F., T.-N.G., Y.-J.C., S.-Y.H., C.-H.Y., C.-C.C.; Validation: C.-T.H., S.-J.F., T.-N.G., Y.-J.C., S.-Y.H.; Formal analysis: C.-T.H., S.-J.F., T.-N.G., Y.-J.C.; Investigation: C.-T.H., S.-J.F., T.-N.G., Y.-J.C., Y.-C.Liao, Y.-C.Lee; Data curation: C.-T.H., S.-J.F., T.-N.G., Y.-J.C.; Visualization: C.-T.H., S.-J.F., T.-N.G.; Writing - original draft: C.-T.H., S.-J.F., T.-N.G., Y.-J.C., C.-H.Y., C.-C.C.; Writing - review & editing: C.-T.H., Y.-C.Liao, S.-Y.H., Y.-C.Lee, C.-H.Y., C.-C.C.; Funding acquisition: C.-T.H., C.-H.Y., C.-C.C.

